# Systematic Benchmarking and Optimization of Retrieval-Augmented Generation (RAG) for Clinical Phenotyping: A Type II Diabetes Case Study in an ICD-Screened Cohort

**DOI:** 10.1101/2025.04.29.25326696

**Authors:** Heekyong Park, Martin Rees, Nils Kruger, Kenshiro Fuse, Victor M. Castro, Vivian Gainer, Nich Wattanasin, Barbara Benoit, Kavishwar B. Wagholikar, Shawn N. Murphy

## Abstract

Traditional EHR phenotyping methods can require substantial disease-specific development and long-term maintenance. While integrating large language models (LLMs) with Retrieval-Augmented Generation (RAG) is promising, the impact of RAG configurations remains underexplored. We systematically evaluated these design choices for type 2 diabetes mellitus (T2DM) phenotyping in an ICD-screened cohort. Using GPT-4o, we processed 170,678 clinical notes from 275 patients. We evaluated seven embedding models and three prompt strategies (baseline, improved zero-shot, and few-shot). Using the improved zero-shot prompt, we tested 350 embedding model–chunk size–retrieval depth combinations on the training set and selected eight optimized configurations for test-set evaluation. Combined with three prompts, these yielded 24 prompt–configuration combinations for comparison with ICD codes and the institutional PheNorm method. RAG achieved a mean sensitivity of 0.9019 (95% CI: 0.8866–0.9172), specificity of 0.7907 (0.7749–0.8066), PPV of 0.9405 (0.9365–0.9445), NPV of 0.6967 (0.6653–0.7281), and F1-score of 0.9204 (0.9124–0.9283). All combinations achieved higher PPV than ICD codes (0.7838). Compared with PheNorm, sensitivity was significantly higher (Holm-adjusted p < 0.001), while specificity differences were not significant. Paired bootstrap analyses showed higher NPV and F1-score, with no significant PPV difference. General-purpose embeddings showed higher sensitivity, whereas biomedical embeddings exhibited higher specificity and PPV. Few-shot prompting increased specificity and PPV but reduced sensitivity. Token usage varied substantially across embedding models. These findings suggest the importance of aligning RAG design with task-specific clinical priorities and token efficiency.

## I. Introduction

ACCURATELY identifying patients with specific clinical conditions or characteristics (i.e., phenotypes) from electronic health records (EHRs) is a fundamental prerequisite for advancing data-driven healthcare and translational medicine. Traditionally, patient cohort identification has relied on structured administrative data (e.g., ICD codes), as well as conventional natural language processing (NLP) and machine learning methods. However, administrative codes are primarily designed for billing purposes and may not accurately reflect a patient’s clinical status. Meanwhile, traditional phenotyping pipelines face challenges in long-term maintenance and scalability. Clinical protocols and medications evolve over time, requiring periodic re-evaluation of existing models, while developing high-performing phenotyping algorithms remains a labor-intensive, disease-specific process.

Recent advances in *Large Language Model (LLM)* technologies have the potential to address these issues, with several studies already reporting the successful use of LLMs to enhance clinical phenotyping [4-9]. Specifically, Retrieval-Augmented Generation (RAG) [11] has emerged as an effective framework for improving LLM performance. By incorporating relevant information retrieved from a local vector database into an LLM’s context, RAG enables the efficient handling of large-scale patient data. It also grounds the model’s response in retrieved patient-specific evidence, which can reduce unsupported outputs. Consequently, recent literature shows the rapid adoption of RAG across the medical field for complex tasks, including patient phenotyping [7], clinical trial eligibility screening [13], decision support [14, 15], clinical Q&A [16, 17], and information extraction [18]. For example, [18] demonstrates that RAG-based approaches produce significant performance improvements over traditional generative methods in clinical summarization.

Despite these advantages, broader computer science literature indicates that RAG performance is highly sensitive to underlying architectural choices, such as the embedding model, text chunk size, and retrieval depth [21-23]. Meanwhile, in the clinical domain, evaluations of these variables remain limited. Existing studies tend to isolate a single factor, such as text segmentation or retrieval depth, or compare a narrow selection of embedding models. Furthermore, these evaluations are frequently conducted on restricted datasets or specific types of clinical notes, rather than the diverse, unstructured records found in real practice. Consequently, rigorous combinatorial benchmarking using real-world medical data is needed to understand how different RAG configurations affect clinical phenotyping.

To address this gap, using T2DM as a specific clinical use case, our study systematically mapped the interrelated effects of core RAG hyperparameters— including chunk sizes, top*-k*, and diverse embedding models. After optimizing the RAG configurations for each embedding model, we evaluated the impact of prompt engineering strategies using GPT-4o. Ultimately, this paper provides an evidence-based evaluation of key design choices for optimizing RAG-based LLM pipelines for clinical phenotyping. Specifically, we benchmark the performance of GPT-4o against administrative ICD code-based phenotyping and an established method, PheNorm [25], an annotation-free algorithm that scores disease phenotypes from ICD and NLP counts adjusted for healthcare utilization.

Furthermore, we investigate how retrieval configuration and prompt engineering jointly influence phenotyping performance and token efficiency. These analyses offer practical insights for selecting RAG configurations for clinical phenotyping.

## II. Methods

### A. Overall Study Design

We evaluated GPT-4o-based T2DM phenotyping using a RAG framework across seven embedding models and three prompt strategies: a baseline prompt without prompt engineering (*P*_*initial*_), an improved zero-shot prompt (*P*_*imprv*_), and a few-shot prompt (P_fewshot_). For each embedding model, chunk size and retrieval depth (i.e., the number of retrieved chunks, top*-k*) were optimized using the training set, and the resulting configurations were evaluated on the test-set against ICD codes and PheNorm. Token usage and RAG configurations were also analyzed (Fig. 1).

**Fig. 1.**
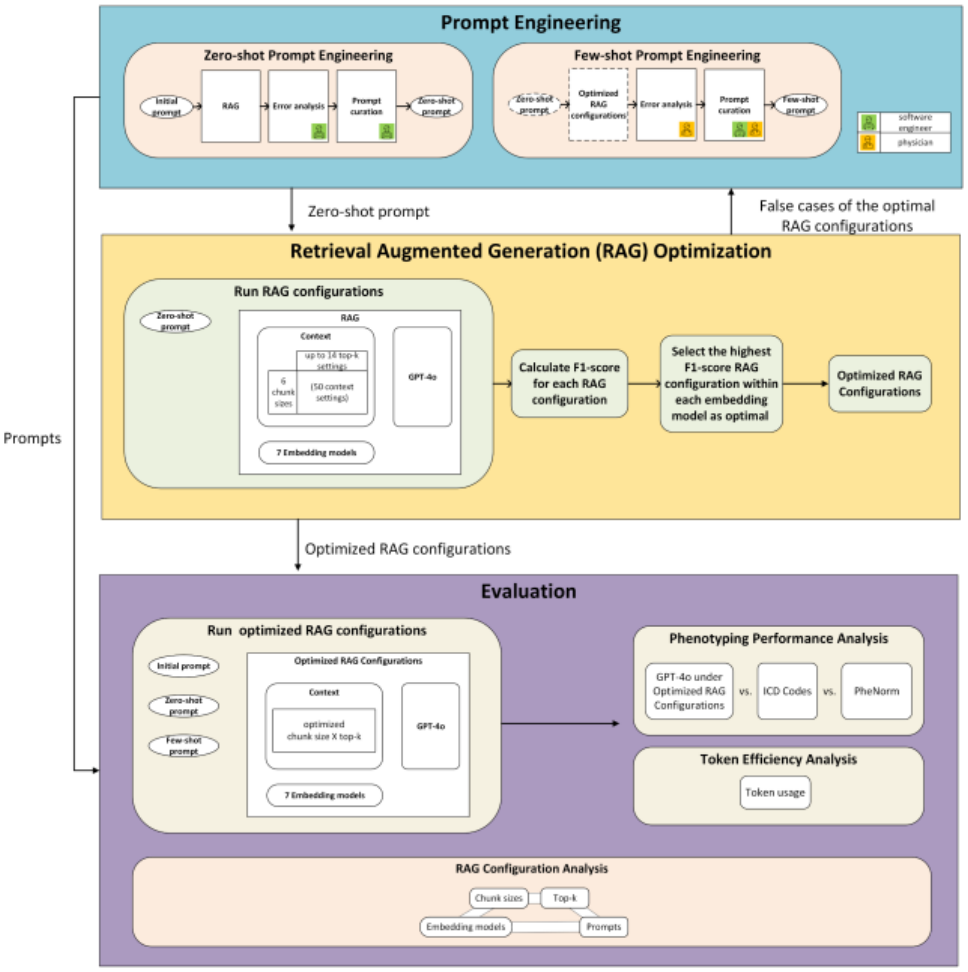
Overall Experimental Design. An improved zero-shot prompt (P_imprv_) was first developed through error analysis of the baseline prompt (P_initial_). Using P_imprv_, 350 combinations of embedding model, chunk size, and retrieval depth (top-k) were evaluated on the training set to optimize RAG configurations. Few-shot prompting (P_fewshot_) was subsequently developed from training-set error analysis. The optimized configurations were then evaluated on the test-set with all three prompt strategies (P_initial_, P_imprv_, and P_fewshot_) against ICD codes and PheNorm. Token efficiency and RAG configuration effects were subsequently analyzed.

### B. Data Description

We reused the T2DM portion of the chart-review labels originally generated to support the retraining of diabetes phenotyping algorithms for the Mass General Brigham (MGB) Biobank Portal. Consistent with phenotyping workflows at MGB, in which ICD codes are first used to identify candidate patients before applying a phenotyping method, 300 patients were randomly selected from the Biobank Portal among those meeting T2DM ICD-code criteria and a data-floor requirement ensuring sufficient clinical text (Appendix A). This staged approach is commonly used because the low prevalence of specific phenotypes in general EHR populations can substantially limit positive predictive value [26].

Each patient was reviewed by a single endocrinologist, who was blinded to the screening criterion by which the patient had been selected and recorded T1DM and T2DM as separate Yes/No determinations. A positive T2DM determination indicated a documented history of T2DM at any point in the record, regardless of current disease status. Free-text comments and structured data elements were also recorded, including diagnosis timing, glycemic and autoimmune laboratory values, and antidiabetic medication exposure.

The T2DM determination served as the gold standard for this study. We excluded 25 patients without a definitive Yes/No label, mainly due to insufficient information in the chart, yielding 275 validated patients, and partitioned them into training (n=90; 67 positive, 23 negative) and test (n=185; 145 positive, 40 negative) sets. The training set was used for RAG configuration optimization and prompt engineering, and the test-set for performance evaluation. We obtained clinical notes for these patients from the MGB Research Patient Data Registry (RPDR) [27, 28] without restriction on note type, spanning each patient’s full available history through the end of the chart-review period. This yielded a corpus of 170,678 unstructured clinical notes.

### C. Retrieval-Augmented Generation (RAG) Framework and Implementation

We implemented a RAG framework to support LLM-based clinical phenotyping. Clinical notes were segmented into fixed-length text chunks using an overlapping sliding window approach. These chunks were vectorized by selected embedding models and stored in a PostgreSQL vector database. Chunks were retrieved via cosine similarity. A preset number of retrieved chunks (top*-k*) or the corresponding parent documents (*pdoc*) were provided as input context to GPT-4o to generate a binary T2DM classification and a structured explanation. For pdoc retrieval, 300-character chunks (hereafter denoted as c300) were used to determine initial relevance. To protect privacy, we used MGB-dedicated Azure OpenAI GPT-4o and text-embedding-3-large, while hosting all other models locally.

### D. Embedding Model Selection

We evaluated seven embedding models (Table I) selected to reflect variations in dimensionality, base architecture, domain specificity (biomedical vs. general), and licensing (open-source vs. commercial). Gte-Qwen2-1.5B-instruct was selected as the top-ranked model on the MTEB leaderboard (overall and retrieval; accessed July 15, 2024) [29] among open-source models with embedding dimensions below 2,000, and text-embedding-3-large was selected as OpenAI’s most recent embedding model at the time of the experiments. Due to PostgreSQL constraints, the maximum embedding dimension was capped at 2,000; accordingly, for text-embedding-3-large, a reduced-dimension version was used. Models were categorized by dimension size: small (512), medium (768), and large (≥1,000).

**TABLE I.** Experimental design: vector embedding models.

| **Embedding model** | **Dimension size** | **Dimension size category** | **Base model** | **(Bio-) medical** | **General** | **Commercial** | **Open source** |
|---|---|---|---|---|---|---|---|
| text-embedding-3-large | 3,072<br>(2,000 after reduction)<sup>*</sup> | large | Unknown |  | v | v |  |
| gte-Qwen2-1.5B-instruct | 1,536 | large | GTE (General Text Embedding) [1] |  | v |  | v |
| GatorTronS [2] | 1,024 | large | BERT [3] | v |  |  | v |
| BiomedBERT [10] | 768 | medium | BERT | v |  |  | v |
| BioBERT [12] | 768 | medium | BERT | v |  |  | v |
| Clinical-Longformer [19] | 768 | medium | Longformer [20] | v |  |  | v |
| Medical-T5-Large | 512 | small | T5 [24] | v |  |  | v |

\* The dimension size of the text-embedding-3-large embedding model is originally 3,072. Due to the dimensional limitation of the PostgreSQL vector database in our RAG system, reduced-dimension version was used.

### E. Iterative Prompt Engineering Process

*P*_*initial*_, consisting of a direct clinical question and structured output instructions, was first evaluated using the text-embedding-3-large model across a limited set of RAG configurations (Table II(a)). False positive (FP) and false negative (FN) cases were reviewed, and recurring error patterns were addressed to iteratively develop P_imprv_. The finalized P_imprv_ was then used for RAG optimization. For P_fewshot_, two physicians independently reviewed P_imprv_ errors from the optimized configurations, categorizing them into retrieval failures or context misinterpretations. Based on these insights, P_fewshot_ was curated by appending five de-identified, clinician-validated text chunks to P_imprv_. Prompt details are summarized in Appendix B.

**TABLE II.**
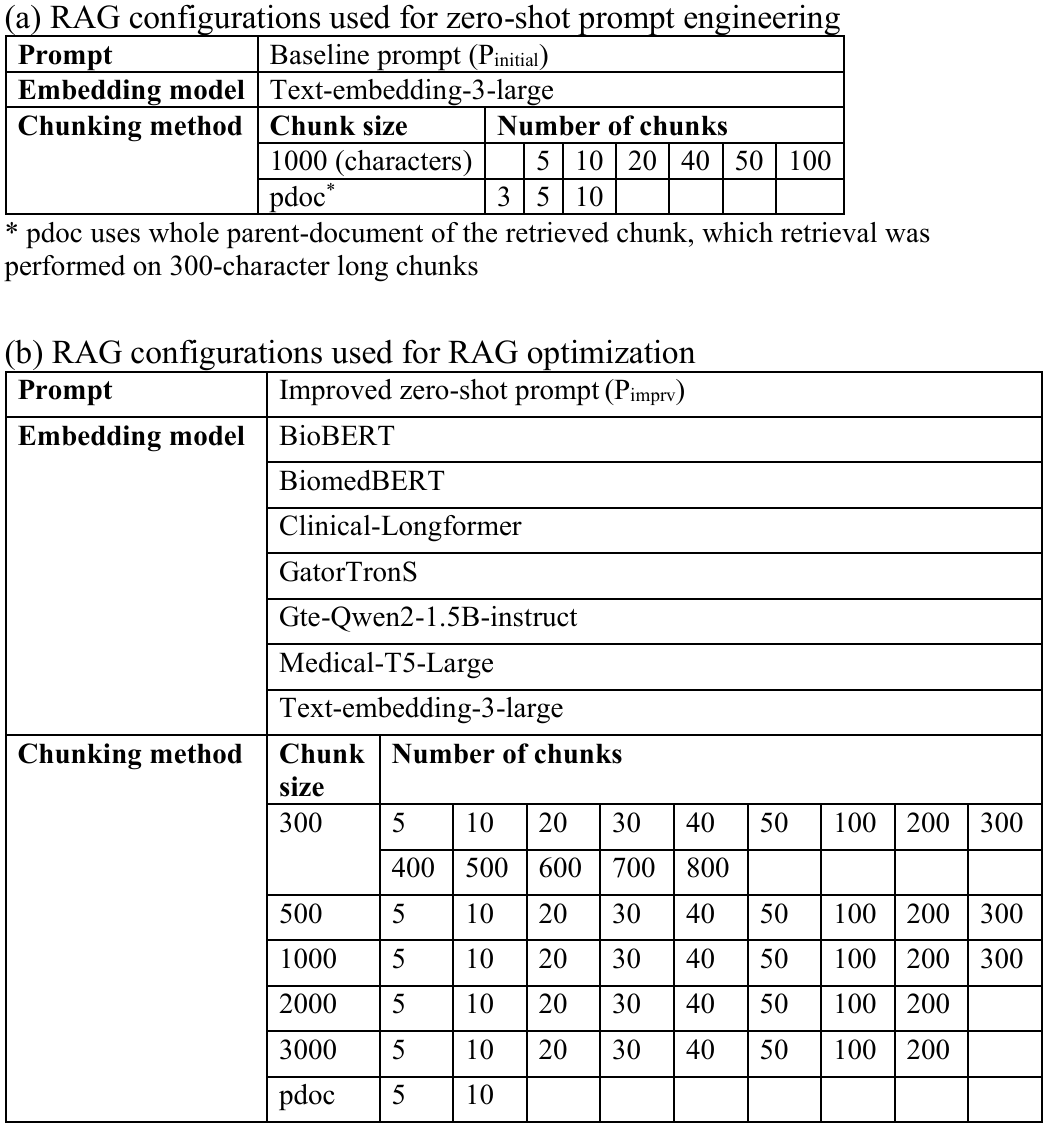
Configurations used for zero-shot prompt engineering and RAG optimization.

| (a) RAG configurations used for zero-shot prompt engineering |  |  |  |  |  |  |  |  |  |  |
|---|---|---|---|---|---|---|---|---|---|---|
| **Prompt** | Baseline prompt (P<sub>initial</sub>) |  |  |  |  |  |  |  |  |  |
| **Embedding model** | Text-embedding-3-large |  |  |  |  |  |  |  |  |  |
| **Chunking method** | **Chunk size** | **Number of chunks** |  |  |  |  |  |  |  |  |
|  | 1000 (characters) |  | 5 | 10 | 20 | 40 | 50 | 100 |  |  |
|  | pdoc<sup>*</sup> | 3 | 5 | 10 |  |  |  |  |  |  |
| (b) RAG configurations used for RAG optimization |  |  |  |  |  |  |  |  |  |  |
| **Prompt** | Improved zero-shot prompt (P<sub>imprv</sub>) |  |  |  |  |  |  |  |  |  |
| **Embedding model** | BioBERT |  |  |  |  |  |  |  |  |  |
|  | BiomedBERT |  |  |  |  |  |  |  |  |  |
|  | Clinical-Longformer |  |  |  |  |  |  |  |  |  |
|  | GatorTronS |  |  |  |  |  |  |  |  |  |
|  | Gte-Qwen2-1.5B-instruct |  |  |  |  |  |  |  |  |  |
|  | Medical-T5-Large |  |  |  |  |  |  |  |  |  |
|  | Text-embedding-3-large |  |  |  |  |  |  |  |  |  |
| **Chunking method** | **Chunk size** | **Number of chunks** |  |  |  |  |  |  |  |  |
|  | 300 | 5 | 10 | 20 | 30 | 40 | 50 | 100 | 200 | 300 |
|  |  | 400 | 500 | 600 | 700 | 800 |  |  |  |  |
|  | 500 | 5 | 10 | 20 | 30 | 40 | 50 | 100 | 200 | 300 |
|  | 1000 | 5 | 10 | 20 | 30 | 40 | 50 | 100 | 200 | 300 |
|  | 2000 | 5 | 10 | 20 | 30 | 40 | 50 | 100 | 200 |  |
|  | 3000 | 5 | 10 | 20 | 30 | 40 | 50 | 100 | 200 |  |
|  | pdoc | 5 | 10 |  |  |  |  |  |  |  |

\* pdoc uses whole parent-document of the retrieved chunk, which retrieval was performed on 300-character long chunks

### F. RAG Hyperparameter Optimization

For each embedding model, using P_imprv_ on the training set, we tested 50 combinations of chunk sizes (c300 to c3000, and pdoc), and top-*k* (Table II(b)). Top-*k* was incrementally increased up to GPT-4o’s context limit. The configuration yielding the highest F1-score, which balances positive predictive value (PPV) and sensitivity, was selected as optimal. If the selected configuration failed for any patient due to context length limitations, the next best-performing configuration without errors was additionally included for that embedding model. Because multiple configurations may yield similar F1-scores, we further evaluated the top five configurations per model to assess variation across different configurations (Section II-G).

### G. Evaluation Metrics, Baseline Comparison, and RAG Configuration Analysis

Phenotyping performance was evaluated on the test-set using the optimized RAG configurations with P_initial_, P_imprv_, and P_fewshot_. When outputs were unavailable for a patient because the input exceeded the GPT-4o API’s maximum input length or triggered the Azure OpenAI content filter, that patient was excluded from both RAG and PheNorm for the corresponding comparison. Overall RAG performance was summarized as the mean of each metric across the 24 prompt-configuration combinations, with 95% CIs computed using the t-distribution across these combinations. PPV was compared with ICD code precision. Sensitivity, specificity, PPV, negative predictive value (NPV), and F1-score were compared with PheNorm from the RPDR, in which patients with a PheNorm score ≥ 0.5 are classified as positive. The RPDR T2DM PheNorm classifications were used as provided, without retraining, recalibration, or threshold adjustment. We used the PheNorm classifications generated in the same calendar month as the endocrinologist chart review. Sensitivity and specificity for each RAG combination were compared with those of PheNorm using exact McNemar tests on paired patient-level predictions, with Holm correction across the 24 comparisons for each metric. Differences in PPV, NPV, and F1-score were evaluated using 95% CIs from 2,000 paired bootstrap resamples of the test-set patients.

We assessed token efficiency via the average retrieved-chunk token count per patient using the GPT-4o tokenizer, excluding prompt instructions, few-shot examples, and output tokens. Furthermore, we conducted a two-stage RAG configuration analysis: 1) evaluating the combined effects of embedding model, chunk size, and top-k using the five highest-F1 configurations per model, and 2) exploring the interplay between prompt strategies and embeddings.

## III. Results

### A. Phenotyping Performance Versus Clinical Baselines

For each of the seven embedding models, the RAG configuration achieving the highest F1-score on the training set was identified as optimal within our evaluation setting. For BiomedBERT, the second-best configuration was also included because the top-ranked setting (pdoc, top-k = 5) exceeded GPT-4o’s context length limit for one patient. In total, eight RAG configurations were selected for test-set evaluation and are summarized in Table III.

**TABLE III.** Optimized RAG Configurations.

| **Embedding model** | **Chunk size** | **Number of chunks** |
|---|---|---|
| BioBERT | 300 | 800 |
| BiomedBERT | 300 | 600 |
| BiomedBERT | pdoc | 5 |
| Clinical-Longformer | pdoc | 10 |
| GatorTronS | 2000 | 200 |
| Gte-Qwen2-1.5B-instruct | 2000 | 20 |
| Medical-T5-Large | 1000 | 200 |
| Text-embedding-3-large | 2000 | 200 |

Table IV summarizes key phenotyping results of RAG compared with ICD codes and PheNorm, and results for all 24 prompt-configuration combinations are provided in Appendix C. Across the 24 evaluated prompt-configuration combinations on the test-set, GPT-4o-based T2DM phenotyping averaged a sensitivity of 0.9019 (95% CI: 0.8866-0.9172), specificity of 0.7907 (95% CI: 0.7749-0.8066), PPV of 0.9405 (95% CI: 0.9365-0.9445), NPV of 0.6967 (95% CI: 0.6653-0.7281), and F1-score of 0.9204 (95% CI: 0.9124-0.9283), with CIs reflecting variability across these combinations.

**TABLE IV.** Phenotyping performance of RAG, ICD codes, and Phenorm.

| **Method / Configuration** | **Sens.** | **Spec.** | **PPV<br>/Precision** | **NPV** | **F1** |
|---|---|---|---|---|---|
| **Clinical Baselines** |  |  |  |  |  |
| ICD codes | - | - | 0.7838 | - | - |
| PheNorm | 0.6000 | 0.9000 | 0.9560 | 0.3830 | 0.7373 |
| **Optimized RAG (GPT-4o)** |  |  |  |  |  |
| Mean of 24 combinations<sup>*</sup> | 0.9019 | 0.7907 | 0.9405 | 0.6967 | 0.9204 |
| BioBERT (c300, k=800), P<sub>imprv</sub><br>(Highest Sens./NPV/F1) | 0.9517 | 0.8000 | 0.9452 | 0.8205 | 0.9485 |
| BiomedBERT (c300, k=600), P<sub>fewshot</sub><br>(Highest Spec./PPV) | 0.8414 | 0.8750 | 0.9606 | 0.6034 | 0.8971 |
| gte-Qwen2-1.5B-instruct (c2000, k=20), P<sub>imprv</sub> (Fewest tokens) | 0.9517 | 0.7750 | 0.9388 | 0.8158 | 0.9452 |

<sup>\*</sup>Mean performance across the eight optimized RAG configurations evaluated with three prompt strategies (P<sub>initial</sub>, P<sub>imprv</sub>, P<sub>fewshot</sub>); see Appendix C for all 24 combinations.

All 24 combinations achieved a PPV exceeding the precision of T2DM ICD codes (0.7838). Compared with PheNorm, RAG operated at a different sensitivity-specificity trade-off: all 24 combinations achieved significantly higher sensitivity (Holm-adjusted p < 0.001), and paired bootstrap analyses showed higher NPV and F1-score, with 95% CIs for the differences excluding zero for all combinations. The maximum improvements in sensitivity, NPV, and F1-score were 0.3517, 0.4375, and 0.2112, respectively. Although specificity was numerically lower across all combinations, none of the differences were statistically significant (Holm-adjusted p ≥ 0.375). Differences in PPV were also small (≤0.0314; 0.9247-0.9606 vs. 0.9560), with all paired bootstrap 95% CIs for the differences including zero. Three combinations exceeded the PPV of PheNorm while maintaining substantially higher sensitivity (0.8414-0.9236 vs. 0.6000).

Fig. 2 visualizes performance and token usage across prompt-embedding model combinations. The highest sensitivity (0.9517) was observed with gte-Qwen2-1.5B-instruct using P_initial_ and P_imprv_, and with BioBERT using P_imprv_. BioBERT with P_imprv_ also achieved the highest NPV (0.8205) and F1-score (0.9485), whereas the highest specificity (0.8750) and PPV (0.9606) were obtained with BiomedBERT (c300, top-k = 600) using P_fewshot_. Among embedding models, gte-Qwen2-1.5B-instruct required the fewest tokens (7,371.85 per patient) while achieving comparable or higher sensitivity, NPV, and F1-score. In contrast, BioBERT required approximately 44,000 tokens, and text-embedding-3-large used the most tokens per patient (73,890.35).

**Fig. 2.**
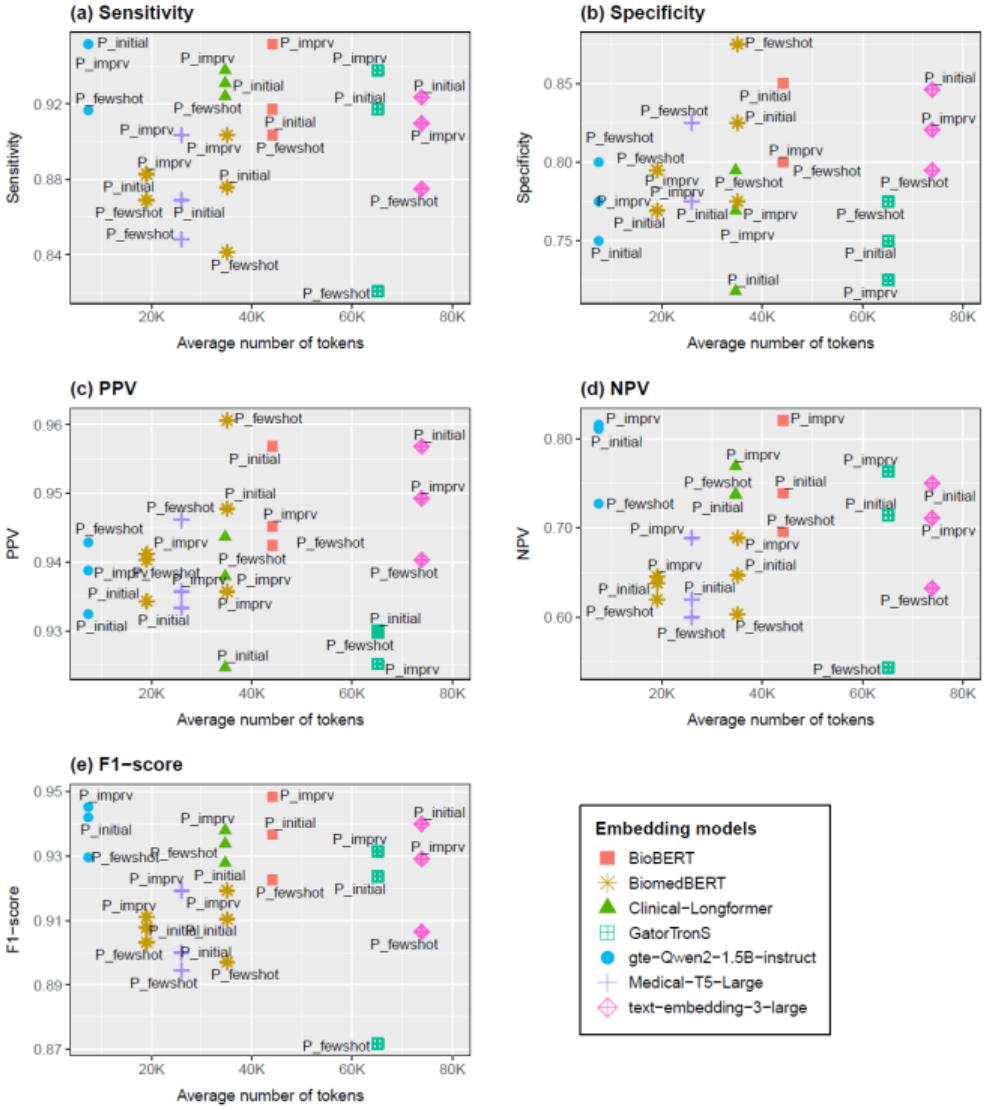
T2DM phenotyping validation results. These are visualizations of optimized RAG’s T2DM phenotyping results validated on test set. The graphs highlight performance and token usage of the test settings. To focus on the differences, each figure uses different y-axis scale. The average number of tokens means average number of tokens used per patient. P_initial, P_imprv, and P_fewshot stand for the baseline prompt, improved zero-shot prompt, and few-shot prompt used in this experiment.

### B. Insights from Error Analysis and Prompt Refinement

#### 1) Insights from Zero-Shot Prompt Engineering

During prompt development, error analysis revealed recurring misinterpretation patterns primarily driven by the LLM’s misalignment with strict clinical phenotyping criteria (e.g., family history or resolved diagnoses) and formatting artifacts in converted clinical text (e.g., bullet-point symbol “?” interpreted as diagnostic uncertainty). Explicit instructions addressing these patterns were incorporated to construct P_imprv_ (Appendix B-2). Retrieval errors also occurred when relevant evidence was excluded at smaller top-k values or became less influential in longer contexts. In one case, the only supporting evidence, ranked 43rd, yielded “YES” with 50 retrieved chunks but “NO” with 100, consistent with the “lost in the middle” phenomenon [30].

#### 2) Insights from Few-Shot Prompt Engineering

Evaluating P_imprv_ using the optimized RAG configurations on the training set yielded 43 FP and FN cases across 18 unique patients. Error analysis classified 3 patients as retrieval failures (observed only with Clinical-Longformer, BioBERT, and BiomedBERT) and 9 as context misinterpretations. The remaining 6 patients did not fall into either category: 2 involved annotation errors, and 4 had ambiguous documentation of diabetes type or status. Context misinterpretations revealed limitations in GPT-4o’s clinical reasoning: the model failed to recognize antidiabetic medications as evidence of T2DM (e.g., liraglutide) or to rule out T2DM despite identifying an alternative diabetes type (e.g., type 1 diabetes).

Other errors occurred when phenotyping required holistic chart review rather than keyword matching. Based on these observations, five clinician-validated examples (two positive, three negative) were selected from the context misinterpretation cases to construct P_fewshot_ (Appendix B-3).

### C. Multi-Dimensional RAG Configuration Analysis and Token Efficiency

Training-set performance and token usage were analyzed for the five highest-F1 configurations per model to examine the combined effects of embedding model, chunk size, and top-k. As shown in Fig. 3, the two large general-purpose embeddings—text-embedding-3-large and gte-Qwen2-1.5B-instruct—consistently achieved the highest sensitivity, NPV, and F1-score. Specifically, these two models accounted for the top ten configurations in sensitivity and NPV and the top eight configurations in F1-score.

**Fig. 3.**
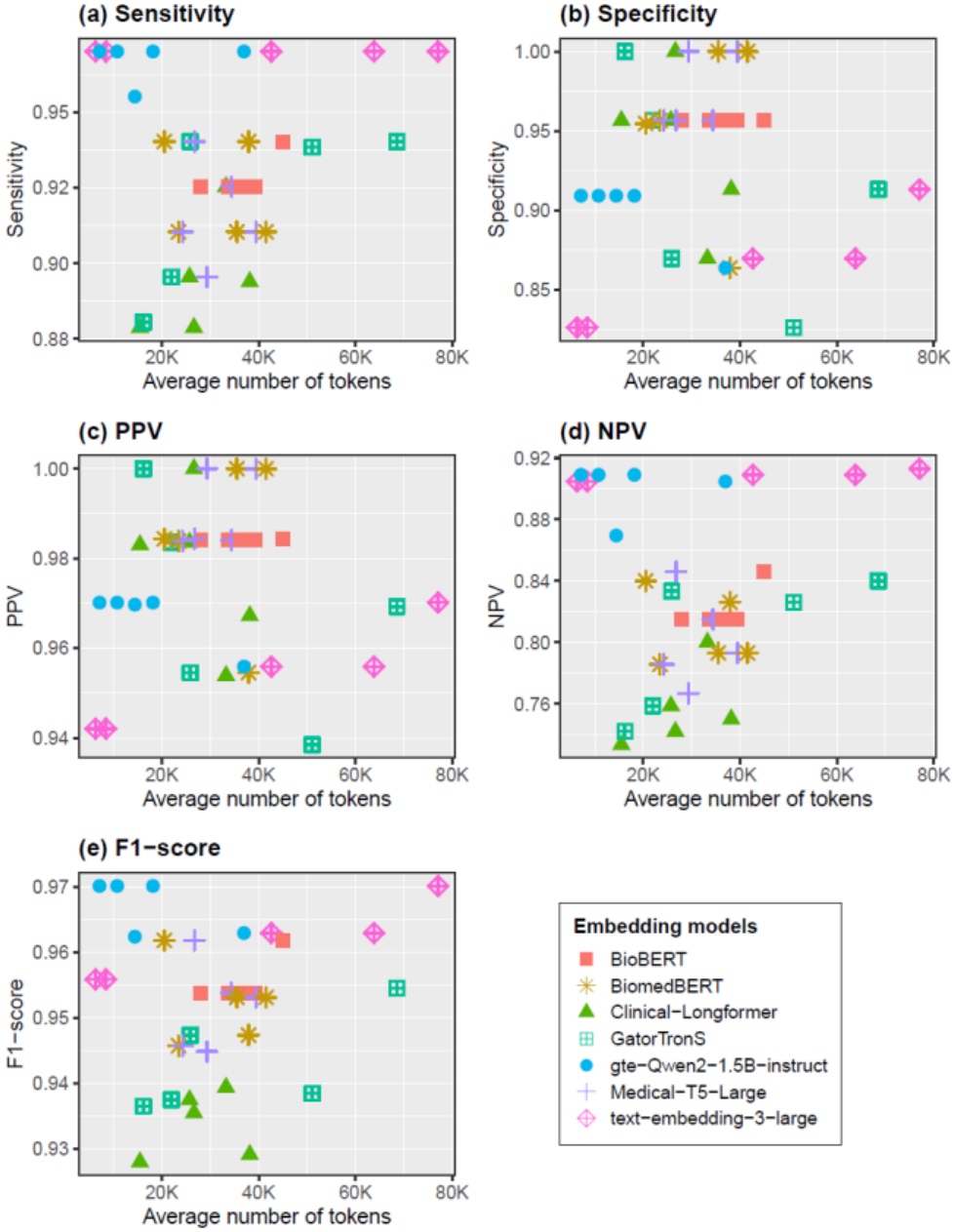
RAG configuration analysis results. Performance and average number of tokens used. The top five F1-scoring configurations for each embedding model were analyzed to assess the combined effects of chunk size, retrieval depth (*top-k*), and embedding choice on performance and the average number of tokens used per patient. Large general-purpose models (text-embedding-3-large, gte-Qwen2-1.5B-instruct) achieved the highest sensitivity, NPV, and F1-scores, while biomedical models excelled in specificity and PPV. Optimal chunk size and top-k varied by model, and token usage of the configurations with the highest F1-score ranged widely, from fewer than 20,000 tokens per patient for gte-Qwen2-1.5B-instruct to over 70,000 for text-embedding-3-large.

In contrast, several biomedical embeddings, including BiomedBERT, Medical-T5-Large, GatorTronS, and Clinical-Longformer, achieved the highest specificity and PPV values, reaching up to 1.0000 in both metrics. The best-performing general embedding (text-embedding-3-large) ranked 20th in specificity (0.9130) and PPV (0.9701). Despite their smaller dimensionality, optimized biomedical embeddings demonstrated competitive performance in F1-score. BioBERT, Medical-T5-Large, and BiomedBERT achieved F1-scores up to 0.9618, within 0.01 of the overall best-performing configuration achieved by general embeddings (0.9701). In contrast, GatorTronS, despite its larger dimensionality, did not outperform these models. Clinical-Longformer exhibited the lowest peak F1-score (0.9394).

Optimal chunk size and top-k varied across embedding models (Fig. 4). Large general-purpose embeddings performed best with larger chunks (c1000 or longer), with all top gte-Qwen2-1.5B-instruct configurations using c2000 chunks. In contrast, biomedical embeddings generally favored smaller chunks (c300) combined with larger top-k (≥300). Some medium-sized models, including BiomedBERT and Clinical-Longformer, achieved high performance using either small chunks or pdoc. No c3000 configuration ranked among the top five.

**Fig. 4.**
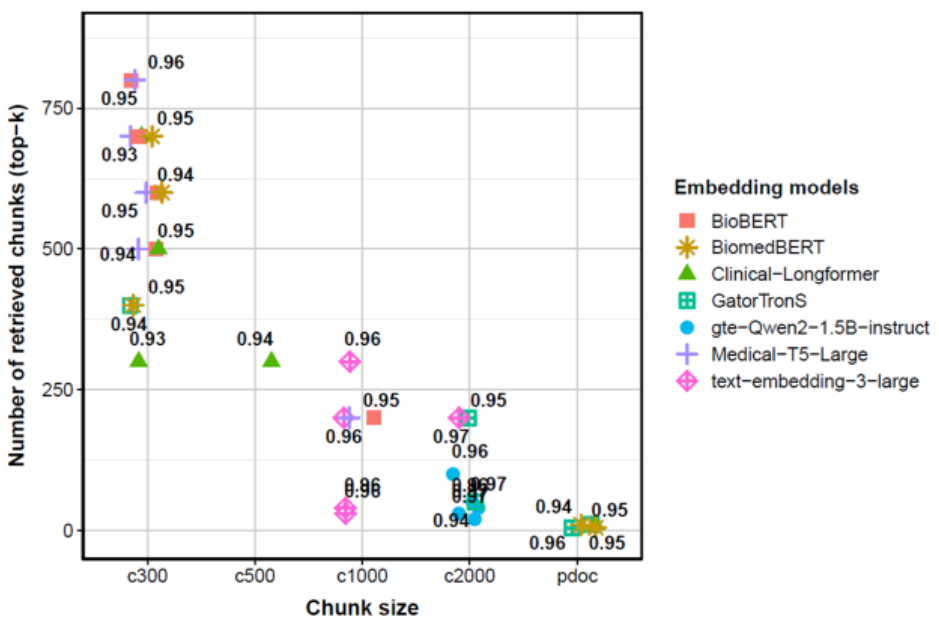
RAG configuration analysis results. top five F1-score RAG configurations of the seven embedding models. This figure illustrates F1-score values of the top five configurations for each embedding model, highlighting how chunk size and the number of retrieved chunks (top-k) influence performance per embedding model. Large general-purpose models performed best with longer chunks (c1000–c2000), while small and medium biomedical models favored short chunks (c300) and higher top-k.

Token efficiency analysis (Fig. 5) showed that small- and medium-sized embeddings operated within a narrow token range, whereas large embeddings exhibited substantial variability. Four of the five highest-F1 configurations for gte-Qwen2-1.5B-instruct used fewer than 20,000 retrieved-chunk tokens per patient, whereas the five highest-F1 configurations for text-embedding-3-large ranged from 6,323.22 to 77,070.05 tokens per patient. At their respective peak-F1 configurations, gte-Qwen2-1.5B-instruct required less than 10% of the tokens used by text-embedding-3-large. For specificity and PPV, GatorTronS achieved the strongest performance with the lowest token usage among all evaluated models. Detailed results are provided in Appendix D.

**Fig. 5.**
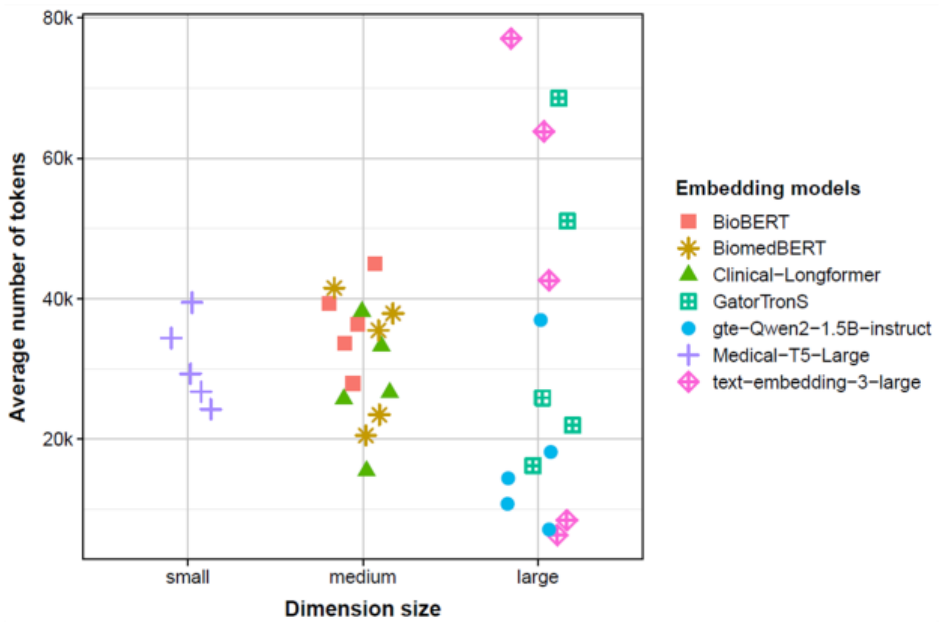
RAG configuration analysis results. F1-score and token usage by embedding model size. This figure highlights token usage across embedding models grouped by embedding model dimension size (small: 512, medium: 768, large: >= 1,000). Small and medium biomedical models exhibited a narrow token range, while large models showed substantial variability.

### D. Impact of Prompt Engineering Strategies

The zero-shot and few-shot prompt strategies exhibited distinct effects on test-set phenotyping performance. As shown in Fig. 2, P_imprv_ increased sensitivity, NPV, and F1-score relative to P_initial_ across most embedding models, with the largest improvement observed in NPV (+0.0814) when paired with BioBERT. However, the impact of P_imprv_ was minimal for gte-Qwen2-1.5B-instruct and BiomedBERT (pdoc, top-k = 5). Improvements in specificity and PPV were limited, with approximately half of the evaluated configurations showing minor gains; Clinical-Longformer was an exception, demonstrating a larger increase in specificity (+0.0513).

In contrast, P_fewshot_ improved specificity and PPV in the majority of configurations but resulted in lower NPV and F1-score compared with P_initial_, except for Clinical-Longformer. No embedding model showed improved sensitivity with P_fewshot_. Clinical-Longformer showed the largest improvement with P_fewshot_, achieving a specificity increase of 0.0770 and representing the only model with concurrent improvements in NPV and F1-score.

## IV. Discussion

We evaluated GPT-4o-based T2DM phenotyping using optimized RAG configurations across seven embedding models and observed strong overall performance. However, our results also demonstrate that naïve application of RAG does not guarantee consistent performance. Across the full RAG design space, substantial variability was observed, with performance gaps within a single embedding model reaching up to 0.7910 in sensitivity for Clinical-Longformer (Appendix E). These findings indicate that phenotyping accuracy depends on the interaction among embedding representations, chunking granularity, and prompt design. Importantly, these findings are grounded in real-world, clinician-validated EHR data drawn from a large, multi-hospital health system.

The RAG configuration analysis revealed differences between general-purpose and biomedical-specific embedding models. General embeddings demonstrated higher sensitivity, NPV, and F1-score, whereas biomedical embeddings achieved higher specificity and PPV. One possible explanation is that the general-purpose embeddings were selected for their strong retrieval performance at the time of the experiments: gte-Qwen2-1.5B-instruct ranked highest on the MTEB leaderboard among open-source models within our dimensionality constraint, and text-embedding-3-large was the most recent embedding model released by OpenAI. Their strong retrieval capability may have contributed to their higher sensitivity. In contrast, biomedical embeddings, trained primarily on biomedical text, may better capture clinical terminology, which can help filter out ambiguous mentions but may miss cases documented mainly through broader context.

Embedding dimensionality may also contribute, although this alone does not fully explain the observed differences, as GatorTronS, the largest biomedical embedding evaluated, did not outperform smaller biomedical models. Biomedical embeddings nevertheless achieved F1-scores comparable to general embeddings when paired with appropriate chunk sizes and top-k, consistent with our prior findings in ischemic stroke phenotyping [31]. Together, these results suggest that embedding selection should be guided by use-case-specific priorities, such as sensitivity versus precision. Although F1-score was used as a balanced optimization criterion in this study, deployment-specific objectives may favor different configurations.

We further observed distinct clusters of optimized hyperparameter settings across embedding models. Larger embeddings generally benefited from longer chunks, whereas small- and medium-sized biomedical embeddings performed well with shorter chunks when sufficient top-k was provided. This suggests that chunk size and top-k should be tuned jointly with the embedding model rather than selected independently. Configurations using c3000 chunks did not rank among top-performing settings, whereas several optimized configurations used full parent documents.

Token efficiency analysis revealed substantial differences in token usage across embedding models. Gte-Qwen2-1.5B-instruct achieved the same peak F1-score as text-embedding-3-large while using less than 10% of the tokens. For specificity and PPV, GatorTronS achieved the highest performance with the lowest token usage among the evaluated models. These findings highlight the importance of jointly considering task-specific performance priorities and token usage when selecting a RAG configuration.

Error analysis and subsequent prompt refinement further indicated that LLM performance depends on explicit task framing and clear phenotyping criteria. P_imprv_, which explicitly addressed EHR formatting artifacts and clarified criteria for historical diagnoses, improved sensitivity, NPV, and F1-score in most configurations. Conversely, while P_fewshot_ enhanced specificity and PPV, it reduced sensitivity and overall F1-score, illustrating the trade-offs inherent in few-shot prompt design [32-35].

This may be attributed to the composition of the few-shot examples, which were selected from difficult misclassified cases and ended with three negative examples after two positive ones (Appendix B-3), potentially encouraging more conservative positive classifications, consistent with previously reported label and recency biases (i.e., a tendency to favor labels appearing more frequently or near the end of the prompt) in few-shot prompting [32]. These findings suggest that a few-shot strategy can help improve cohort precision, at the cost of reduced sensitivity. In addition, key evidence was sometimes ranked low during retrieval and excluded from the model context, suggesting that improved retrieval and ranking strategies may further enhance RAG performance.

Overall, this study demonstrates the potential of LLM-based RAG systems to support clinical phenotyping while highlighting the importance of careful prompt design, retrieval configuration, and embedding selection. Limitations include the evaluation of a single LLM and a single disease domain, as well as the dimensionality constraint of our PostgreSQL vector database, which required using a reduced-dimension version of text-embedding-3-large and may have affected its performance. Additionally, because our cohort was sampled after ICD-based screening to reflect institutional two-stage phenotyping workflows, the reported PPV and NPV reflect performance in an enriched population rather than an unscreened, population-wide EHR cohort. Future work will extend this framework to additional phenotypes, LLMs, and retrieval strategies to further characterize the generalizability of these findings.

## V. Conclusion

This study demonstrates that LLM-based phenotyping using RAG achieves strong performance for T2DM identification through systematic optimization. Our extensive evaluation of embedding models, retrieval parameters, and prompt strategies shows that RAG performance is highly sensitive to design choices, and naïve application can result in substantial variability. Optimized GPT-4o RAG configurations achieved higher PPV than ICD codes and higher sensitivity, NPV, and F1-score than PheNorm, with no significant difference in PPV or specificity.

Our results reveal clear trade-offs across embedding models and prompt strategies. General-purpose embeddings achieved higher sensitivity, whereas biomedical embeddings demonstrated higher PPV under optimized retrieval settings. In addition, improved zero-shot prompt engineering increased sensitivity, NPV, and F1-score, whereas few-shot prompt engineering improved specificity and PPV, with corresponding reductions in sensitivity. Token usage also varied substantially across embedding models. Together, these findings highlight the importance of considering task-specific performance priorities and token efficiency when selecting and optimizing RAG configurations.

Overall, these findings provide practical guidance for designing RAG pipelines for clinical phenotyping. Although focused on a single disease domain and LLM, the observed configuration-dependent patterns may extend to other phenotyping tasks. Future work will extend RAG evaluation to additional phenotypes to support the integration of LLM technology for institutional phenotyping pipelines.

## Data Availability

All shareable data produced in the present work are contained in the manuscript.

