## Appendix for "Systematic Benchmarking and Optimization of Retrieval-Augmented Generation (RAG) for Clinical Phenotyping: A Type II Diabetes Case Study in an ICD-Screened Cohort"

### Appendix A. Patient selection

#### Patient selection method

Three hundred patients were randomly selected from the Mass General Brigham Biobank Portal[1] among those meeting all of the following criteria:

- The patient has at least one Type II diabetes mellitus (T2DM) diagnosis code (Table 1)
- The patient meets the following data floor to ensure a minimum amount of records
  - Has at least 3 diagnosis codes greater than 30 days apart
  - Has at least 1 diagnosis code after 1/1/2005
  - Has more than 1 clinical note

**Table 1. T2DM ICD codes used to screen patients**

| **ICD version** | **T2DM Codes** |
|---|---|
| ICD-9-CM | 250.X0, 250.X2* |
| ICD-10-CM | E11.X, E11.XX, E11.XXX, E11.XXXX* |

*X stands for any number from 0 to 9.

### Appendix B. Prompts

#### 1. Initial prompt ($P_{initial}$)

| Instruction type | Prompt content |
|---|---|
| Question | Has the patient been diagnosed with type 2 diabetes? |
| Output format | Answer YES or NO. JSON formatted response: {“Answer” : “YES \| NO”, “Reason”: “Reason Here”} |

#### 2. Improved zero-shot prompt ($P_{imprv}$)

| Instruction type | Prompt content |
|---|---|
| Task (Question) | Has the patient been diagnosed with type 2 diabetes? |
| Output format | Answer YES or NO. JSON formatted response: {“Answer”: “YES \| NO”, “Reason”: “Reason Here”} |
| Goal | This task is for patient phenotyping, using the patient's EHR clinical notes, to build type 2 diabetes (t2dm) patient cohort for clinical research. |
| Instructions for improved decision (Detailed task instructions) | Answer “YES” if there is any mention or indication that the patient has been diagnosed with or treated for 't2dm'. Consider any term related to 't2dm' including synonyms or related medical terminology, and look for explicit mentions or implications of a diagnosis or treatment. |
| Instructions to avoid loose-interpretation | The identification of the disease should be precise. If there the patient does not have trivial evidence of 't2dm' phenotype, answer “NO”.<br>Do not conclude the following cases as “YES”: the other types of diabetes or diabetes mellitus without specific evidence of type 2. |
| Overall instruction to consider clinical notes' structures | The provided texts are part of real EHR patient notes. Consider incorporating clinical note's structural information, such as section title (problem list, History of Present Illness, Diagnoses, etc.), for identifying 't2dm' |
| - Bullet-pointed list | Some text parts contain bullet-pointed list converted from PDF files, analyze the content considering the unique formatting. Notably, some terms or phrases might be split across multiple lines or separated by multiple spaces, reflecting the original table cell boundaries and alignments.<br>Therefore, special characters, such as ”?” mark right in front of<br>a phrase or a medical concept may indicate a bullet point, not the semantic m<br>eaning such as ”questionable” or ”uncertain”. |
| - Table | Some text parts originally formatted as tables in PDF, analyze the content considering the unique formatting. There can be multiple lines of table content text delimited by consecutive spaces to fill the space until the next cell starts. Consider header information when a tabular content is recognized. Some phrase or a term inside the table may appear split in 2 lines, reflecting line change due to space limitation in a table cell. The headers converted from these tables still appear in the text, influencing how data should be read and associated. Use the headers as guides to understand the context and relationship between different pieces of data. |

#### 3. Few-shot prompt ($P_{fewshot}$)

(a) Prompt structure

| Instruction type | Prompt content |
|---|---|
| Task and instructions | Same as $P_{imprv}$ |
| Few-shot examples | Here are examples of Context and Answer:<br>[Example 1]<br>Context 1: "*selected chunk text*"<br>Answer 1: "YES"<br><br>[Example 2]<br>Context 2: "*selected chunk text*"<br>Answer 2: "YES"<br><br>[Example 3]<br>Context 3-1: "*selected chunk text*"<br>Context 3-2: "*selected chunk text*"<br>Answer 3: "NO"<br><br>[Example 4]<br>Context 4: "*selected chunk text*"<br>Answer 4: "NO"<br><br>[Example 5]<br>Context 5: "*selected chunk text*"<br>Answer 5: "NO" |

(b) Summary of few-shot examples used in $P_{fewshot}$

| Example | Summary of context | Answer | Purpose |
|---|---|---|---|
| 1 | Home use of basal insulin and a GLP-1 receptor agonist | YES | Recognize a treatment pattern suggestive of T2DM |
| 2 | Basal insulin and a GLP-1 receptor agonist in a medication list | YES | Recognize a treatment pattern suggestive of T2DM |
| 3 | Insulin-treated diabetes in an assessment and plan, with type 1 diabetes documented in the medical history | NO | Distinguish other types of diabetes from T2DM |
| 4 | Cystic fibrosis–related diabetes previously treated with insulin and currently managed with diet, with prior partial pancreatectomy | NO | Distinguish other types of diabetes from T2DM |
| 5 | Steroid-induced diabetes in a patient receiving long-term corticosteroid therapy for vasculitis | NO | Distinguish other types of diabetes from T2DM |

* Examples were selected from misclassified cases identified during error analysis of the training set and reviewed by clinicians. Contexts are summarized to protect patient privacy.

#### Appendix C. Performance of T2DM identification by ICD codes, PheNorm phenotyping algorithm, and GPT-4o with the optimized RAG configurations on the test set

| Phenotyping method | | | | | | Sensitivity | Specificity | PPV / Precision | NPV | F1 | Avg. num tokens |
|---|---|---|---|---|---|---|---|---|---|---|---|
| ICD codes | | | | | | - | - | 0.7838 | - | - | |
| PheNorm | | | | | | 0.6000 | 0.9000 | 0.9560 | 0.3830 | 0.7373 | |
| GPT-4o | Mean (95% CI)* | | | | | 0.9019 (0.8866-0.9172) | 0.7907 (0.7749-0.8066) | 0.9405 (0.9365-0.9445) | 0.6967 (0.6653-0.7281) | 0.9204 (0.9124-0.9283) | - |
| | Prompt | | Embedding model | Chunk size | Num chunks | | | | | | |
| | $P_{initial}$ | | BioBERT | 300 | 800 | 0.9172 | 0.8500 | 0.9568 | 0.7391 | 0.9366 | 44,131.36 |
| | | | BiomedBERT | 300 | 600 | 0.8759 | 0.8250 | 0.9478 | 0.6471 | 0.9104 | 35,062.25 |
| | | | BiomedBERT | pdoc | 5 | 0.8828 | 0.7692 | 0.9343 | 0.6383 | 0.9078 | 19,025.18 |
| | | | Clinical-Longformer | pdoc | 10 | 0.9310 | 0.7179 | 0.9247 | 0.7368 | 0.9278 | 34,682.29 |
| | | | GatorTronS | 2000 | 200 | 0.9172 | 0.7500 | 0.9301 | 0.7143 | 0.9236 | 65,111.79 |
| | | | gte-Qwen2-1.5B-instruct | 2000 | 20 | 0.9517 | 0.7500 | 0.9324 | 0.8108 | 0.9420 | 7,371.85 |
| | | | Medical-T5-Large | 1000 | 200 | 0.8690 | 0.7750 | 0.9333 | 0.6200 | 0.9000 | 25,967.30 |
| | | | text-embedding-3-large | 2000 | 200 | 0.9236 | 0.8462 | 0.9568 | 0.7500 | 0.9399 | 73,890.35 |
| | $P_{imprv}$ | | BioBERT | 300 | 800 | 0.9517 | 0.8000 | 0.9452 | 0.8205 | 0.9485 | 44,131.36 |
| | | | BiomedBERT | 300 | 600 | 0.9034 | 0.7750 | 0.9357 | 0.6889 | 0.9193 | 35,062.25 |
| | | | BiomedBERT | pdoc | 5 | 0.8828 | 0.7949 | 0.9412 | 0.6458 | 0.9110 | 19,025.18 |
| | | | Clinical-Longformer | pdoc | 10 | 0.9379 | 0.7692 | 0.9379 | 0.7692 | 0.9379 | 34,682.29 |
| | | | GatorTronS | 2000 | 200 | 0.9379 | 0.7250 | 0.9252 | 0.7632 | 0.9315 | 65,111.79 |
| | | | gte-Qwen2-1.5B-instruct | 2000 | 20 | 0.9517 | 0.7750 | 0.9388 | 0.8158 | 0.9452 | 7,371.85 |
| | | | Medical-T5-Large | 1000 | 200 | 0.9034 | 0.7750 | 0.9357 | 0.6889 | 0.9193 | 25,967.30 |
| | | | text-embedding-3-large | 2000 | 200 | 0.9097 | 0.8205 | 0.9493 | 0.7111 | 0.9291 | 73,890.35 |
| | $P_{fewshot}$ | | BioBERT | 300 | 800 | 0.9034 | 0.8000 | 0.9424 | 0.6957 | 0.9225 | 44,131.36 |
| | | | BiomedBERT | 300 | 600 | 0.8414 | 0.8750 | 0.9606 | 0.6034 | 0.8971 | 35,062.25 |
| | | | BiomedBERT | pdoc | 5 | 0.8690 | 0.7949 | 0.9403 | 0.6200 | 0.9032 | 19,025.18 |
| | | | Clinical-Longformer | pdoc | 10 | 0.9241 | 0.7949 | 0.9437 | 0.7381 | 0.9338 | 34,682.29 |
| | | | GatorTronS | 2000 | 200 | 0.8207 | 0.7750 | 0.9297 | 0.5439 | 0.8718 | 65,111.79 |

| | | | | | | | | | | |
|---|---|---|---|---|---|---|---|---|---|---|
| | | gte-Qwen2-1.5B-instruct | 2000 | 20 | 0.9167 | 0.8000 | 0.9429 | 0.7273 | 0.9296 | 7,371.85 |
| | | Medical-T5-Large | 1000 | 200 | 0.8483 | 0.8250 | 0.9462 | 0.6000 | 0.8945 | 25,967.30 |
| | | text-embedding-3-large | 2000 | 200 | 0.8750 | 0.7949 | 0.9403 | 0.6327 | 0.9065 | 73,890.35 |

* Mean and 95% confidence interval across the 24 prompt–configuration combinations (eight optimized configurations across three prompts) listed below.

#### Appendix D. Secondary RAG configuration analysis results: results of top five F1-score configurations on the training set

This table describes secondary RAG configuration experimental results, which are selected by five highest F1-score results of each tested embedding model.

| Embedding model | Dimension | Dim. category | Chunk size | Num chunks | Avg. num tokens per patient | Sensitivity | Specificity | PPV | NPV | F1 |
|---|---|---|---|---|---|---|---|---|---|---|
| BioBERT | 768 | medium | 300 | 500 | 27926.87 | 0.9254 | 0.9565 | 0.9841 | 0.8148 | 0.9538 |
| | | | 300 | 600 | 33643.51 | 0.9254 | 0.9565 | 0.9841 | 0.8148 | 0.9538 |
| | | | 300 | 700 | 39363.18 | 0.9254 | 0.9565 | 0.9841 | 0.8148 | 0.9538 |
| | | | 300 | 800 | 44996.34 | 0.9403 | 0.9565 | 0.9844 | 0.8462 | 0.9618 |
| | | | 1000 | 200 | 36336.22 | 0.9254 | 0.9565 | 0.9841 | 0.8148 | 0.9538 |
| BiomedBERT | 768 | medium | 300 | 400 | 23463.41 | 0.9104 | 0.9565 | 0.9839 | 0.7857 | 0.9457 |
| | | | 300 | 600 | 35503.87 | 0.9104 | 1.0000 | 1.0000 | 0.7931 | 0.9531 |
| | | | 300 | 700 | 41527.22 | 0.9104 | 1.0000 | 1.0000 | 0.7931 | 0.9531 |
| | | | pdoc | 5 | 20512.05 | 0.9403 | 0.9545 | 0.9844 | 0.8400 | 0.9618 |
| | | | pdoc | 10 | 37920.24 | 0.9403 | 0.8636 | 0.9545 | 0.8261 | 0.9474 |
| Clinical-Longformer | 768 | medium | 300 | 300 | 15484.71 | 0.8788 | 0.9565 | 0.9831 | 0.7333 | 0.9280 |
| | | | 300 | 500 | 26638.08 | 0.8788 | 1.0000 | 1.0000 | 0.7419 | 0.9355 |
| | | | 300 | 700 | 38172.99 | 0.8939 | 0.9130 | 0.9672 | 0.7500 | 0.9291 |
| | | | 500 | 300 | 25721.04 | 0.8955 | 0.9565 | 0.9836 | 0.7586 | 0.9375 |
| | | | pdoc | 10 | 33268.39 | 0.9254 | 0.8696 | 0.9538 | 0.8000 | 0.9394 |
| GatorTronS | 1024 | large | 300 | 400 | 21981.61 | 0.8955 | 0.9565 | 0.9836 | 0.7586 | 0.9375 |
| | | | 2000 | 50 | 16188.38 | 0.8806 | 1.0000 | 1.0000 | 0.7419 | 0.9365 |
| | | | 2000 | 200 | 68568.66 | 0.9403 | 0.9130 | 0.9692 | 0.8400 | 0.9545 |
| | | | pdoc | 5 | 25883.82 | 0.9403 | 0.8696 | 0.9545 | 0.8333 | 0.9474 |
| | | | pdoc | 10 | 51061.01 | 0.9385 | 0.8261 | 0.9385 | 0.8261 | 0.9385 |
| gte-Qwen2-1.5B-instruct | 1536 | large | 2000 | 20 | 7129.71 | 0.9701 | 0.9091 | 0.9701 | 0.9091 | 0.9701 |
| | | | 2000 | 30 | 10772.46 | 0.9701 | 0.9091 | 0.9701 | 0.9091 | 0.9701 |
| | | | 2000 | 40 | 14425.85 | 0.9552 | 0.9091 | 0.9697 | 0.8696 | 0.9624 |

| | | | | | | | | | |
|---|---|---|---|---|---|---|---|---|---|
| | | | 2000 | 50 | 18167.26 | 0.9701 | 0.9091 | 0.9701 | 0.9091 | 0.9701 |
| | | | 2000 | 100 | 36967.94 | 0.9701 | 0.8636 | 0.9559 | 0.9048 | 0.9630 |
| Medical-T5-Large | 512 | small | 300 | 500 | 24270.56 | 0.9104 | 0.9565 | 0.9839 | 0.7857 | 0.9457 |
| | | | 300 | 600 | 29322.37 | 0.8955 | 1.0000 | 1.0000 | 0.7667 | 0.9449 |
| | | | 300 | 700 | 34387.11 | 0.9254 | 0.9565 | 0.9841 | 0.8148 | 0.9538 |
| | | | 300 | 800 | 39453.08 | 0.9104 | 1.0000 | 1.0000 | 0.7931 | 0.9531 |
| | | | 1000 | 200 | 26758.58 | 0.9403 | 0.9565 | 0.9844 | 0.8462 | 0.9618 |
| text-embedding-3-large | 2,000* | large | 1000 | 30 | 6323.22 | 0.9701 | 0.8261 | 0.9420 | 0.9048 | 0.9559 |
| | | | 1000 | 40 | 8445.69 | 0.9701 | 0.8261 | 0.9420 | 0.9048 | 0.9559 |
| | | | 1000 | 200 | 42627.19 | 0.9701 | 0.8696 | 0.9559 | 0.9091 | 0.9630 |
| | | | 1000 | 300 | 63832.84 | 0.9701 | 0.8696 | 0.9559 | 0.9091 | 0.9630 |
| | | | 2000 | 200 | 77070.05 | 0.9701 | 0.9130 | 0.9701 | 0.9130 | 0.9701 |

* The dimension size of the text-embedding-3-large embedding model is originally 3,072. Due to the dimensional limitation of the PostgreSQL vector database in our RAG system, reduced-dimension version was used.

#### Appendix E. Performance of each embedding model across all configurations used in the RAG optimization process on the training set

| Embedding_model | n | eval_metric | mean | 95% CI | min | max |
|---|---|---|---|---|---|---|
| BioBERT | 50 | Sensitivity | 0.7557 | 0.7118-0.7996 | 0.2836 | 0.9403 |
| | | Specificity | 0.9416 | 0.9305-0.9527 | 0.8261 | 1.0000 |
| | | NPV | 0.6077 | 0.5683-0.6471 | 0.3239 | 0.8462 |
| | | PPV | 0.9746 | 0.9700-0.9792 | 0.9375 | 1.0000 |
| | | F1 | 0.8409 | 0.8089-0.8729 | 0.4419 | 0.9618 |
| BiomedBERT | 50 | Sensitivity | 0.7877 | 0.7481-0.8273 | 0.3731 | 0.9403 |
| | | Specificity | 0.9456 | 0.9322-0.9589 | 0.8261 | 1.0000 |
| | | NPV | 0.6413 | 0.6037-0.6790 | 0.3538 | 0.8400 |
| | | PPV | 0.9784 | 0.9733-0.9834 | 0.9365 | 1.0000 |
| | | F1 | 0.8643 | 0.8374-0.8911 | 0.5435 | 0.9618 |
| Clinical-Longformer | 50 | Sensitivity | 0.6153 | 0.5517-0.6788 | 0.1343 | 0.9254 |
| | | Specificity | 0.9537 | 0.9377-0.9696 | 0.8261 | 1.0000 |
| | | NPV | 0.5077 | 0.4660-0.5494 | 0.2750 | 0.8000 |
| | | PPV | 0.9767 | 0.9691-0.9842 | 0.9000 | 1.0000 |
| | | F1 | 0.7266 | 0.6720-0.7813 | 0.2338 | 0.9394 |
| GatorTronS | 50 | Sensitivity | 0.8354 | 0.8162-0.8546 | 0.6418 | 0.9403 |
| | | Specificity | 0.9345 | 0.9243-0.9446 | 0.8261 | 1.0000 |
| | | NPV | 0.6728 | 0.6495-0.6962 | 0.4773 | 0.8400 |
| | | PPV | 0.9743 | 0.9705-0.9780 | 0.9385 | 1.0000 |
| | | F1 | 0.8978 | 0.8864-0.9092 | 0.7748 | 0.9545 |
| gte-Qwen2-1.5B-instruct | 50 | Sensitivity | 0.8886 | 0.8670-0.9102 | 0.5821 | 0.9701 |
| | | Specificity | 0.8766 | 0.8584-0.8948 | 0.6364 | 1.0000 |
| | | NPV | 0.7502 | 0.7198-0.7807 | 0.4167 | 0.9091 |
| | | PPV | 0.9555 | 0.9496-0.9613 | 0.8873 | 1.0000 |
| | | F1 | 0.9188 | 0.9061-0.9315 | 0.7156 | 0.9701 |

| | | | | | | |
|---|---|---|---|---|---|---|
| Medical-T5-Large | 50 | Sensitivity | 0.6328 | 0.5644-0.7011 | 0.1493 | 0.9403 |
| | | Specificity | 0.9738 | 0.9650-0.9826 | 0.8696 | 1.0000 |
| | | NPV | 0.5363 | 0.4871-0.5855 | 0.2875 | 0.8462 |
| | | PPV | 0.9874 | 0.9830-0.9918 | 0.9286 | 1.0000 |
| | | F1 | 0.7399 | 0.6820-0.7977 | 0.2597 | 0.9618 |
| text-embedding-3-large | 50 | Sensitivity | 0.9231 | 0.9044-0.9419 | 0.6119 | 0.9706 |
| | | Specificity | 0.8119 | 0.7933-0.8305 | 0.6957 | 0.9565 |
| | | NPV | 0.8033 | 0.7750-0.8316 | 0.4468 | 0.9130 |
| | | PPV | 0.9354 | 0.9296-0.9413 | 0.8986 | 0.9796 |
| | | F1 | 0.9276 | 0.9170-0.9382 | 0.7455 | 0.9701 |

#### Appendix F. Statistical comparison of T2DM phenotyping performance between RAG and PheNorm on the test set

| Prompt | Configuration | n | Sens. Holm *p* | Spec. Holm *p* | ΔPPV (95% CI) | ΔNPV (95% CI) | ΔF1 (95% CI) |
|---|---|---|---|---|---|---|---|
| $P_{initial}$ | BioBERT (c300, k800) | 185 | <0.001 | 1 | 0.0008 (-0.0460, 0.0515) | 0.3562 (0.2449, 0.4651) | 0.1993 (0.1395, 0.2660) |
| | BiomedBERT (c300, k600) | 185 | <0.001 | 1 | -0.0083 (-0.0548, 0.0357) | 0.2641 (0.1635, 0.3735) | 0.1731 (0.1090, 0.2418) |
| | BiomedBERT (pdoc, k5) | 184 | <0.001 | 1 | -0.0217 (-0.0653, 0.0211) | 0.2620 (0.1625, 0.3712) | 0.1705 (0.1140, 0.2331) |
| | Clinical-Longformer (pdoc, k10) | 184 | <0.001 | 0.375 | -0.0314 (-0.0719, 0.0045) | 0.3605 (0.2451, 0.4840) | 0.1906 (0.1335, 0.2544) |
| | GatorTronS (c2000, k200) | 185 | <0.001 | 1 | -0.0260 (-0.0733, 0.0182) | 0.3313 (0.2201, 0.4498) | 0.1863 (0.1225, 0.2510) |
| | gte-Qwen2-1.5B-instruct (c2000, k20) | 185 | <0.001 | 1 | -0.0236 (-0.0719, 0.0233) | 0.4278 (0.3142, 0.5569) | 0.2047 (0.1426, 0.2688) |
| | Medical-T5-Large (c1000, k200) | 185 | <0.001 | 1 | -0.0227 (-0.0692, 0.0191) | 0.2370 (0.1390, 0.3507) | 0.1627 (0.1035, 0.2305) |
| | text-embedding-3-large (c2000, k200) | 183 | <0.001 | 1 | 0.0008 (-0.0412, 0.0442) | 0.3696 (0.2629, 0.4828) | 0.1995 (0.1414, 0.2614) |
| $P_{imprv}$ | BioBERT (c300, k800) | 185 | <0.001 | 1 | -0.0108 (-0.0556, 0.0349) | 0.4375 (0.3261, 0.5581) | 0.2112 (0.1522, 0.2772) |
| | BiomedBERT (c300, k600) | 185 | <0.001 | 1 | -0.0203 (-0.0689, 0.0257) | 0.3059 (0.1932, 0.4196) | 0.1820 (0.1184, 0.2479) |
| | BiomedBERT (pdoc, k5) | 184 | <0.001 | 1 | -0.0149 (-0.0609, 0.0294) | 0.2695 (0.1731, 0.3792) | 0.1737 (0.1152, 0.2359) |
| | Clinical-Longformer (pdoc, k10) | 184 | <0.001 | 1 | -0.0181 (-0.0609, 0.0232) | 0.3929 (0.2804, 0.5158) | 0.2006 (0.1370, 0.2705) |
| | GatorTronS (c2000, k200) | 185 | <0.001 | 1 | -0.0309 (-0.0795, 0.0190) | 0.3802 (0.2599, 0.5067) | 0.1942 (0.1284, 0.2627) |
| | gte-Qwen2-1.5B-instruct (c2000, k20) | 185 | <0.001 | 1 | -0.0173 (-0.0633, 0.0297) | 0.4328 (0.3134, 0.5562) | 0.2079 (0.1462, 0.2740) |
| | Medical-T5-Large (c1000, k200) | 185 | <0.001 | 1 | -0.0203 (-0.0656, 0.0258) | 0.3059 (0.1929, 0.4314) | 0.1820 (0.1177, 0.2551) |
| | text-embedding-3-large (c2000, k200) | 183 | <0.001 | 1 | -0.0068 (-0.0498, 0.0368) | 0.3307 (0.2214, 0.4475) | 0.1887 (0.1284, 0.2524) |
| $P_{fewshot}$ | BioBERT (c300, k800) | 185 | <0.001 | 1 | -0.0136 (-0.0597, 0.0317) | 0.3127 (0.2013, 0.4314) | 0.1853 (0.1230, 0.2551) |
| | BiomedBERT (c300, k600) | 185 | <0.001 | 1 | 0.0046 (-0.0403, 0.0536) | 0.2205 (0.1193, 0.3305) | 0.1598 (0.0955, 0.2278) |
| | BiomedBERT (pdoc, k5) | 184 | <0.001 | 1 | -0.0157 (-0.0614, 0.0297) | 0.2437 (0.1443, 0.3529) | 0.1659 (0.1079, 0.2306) |
| | Clinical-Longformer (pdoc, k10) | 184 | <0.001 | 1 | -0.0124 (-0.0514, 0.0259) | 0.3618 (0.2542, 0.4910) | 0.1965 (0.1355, 0.2631) |
| | GatorTronS (c2000, k200) | 185 | <0.001 | 1 | -0.0264 (-0.0763, 0.0238) | 0.1609 (0.0694, 0.2594) | 0.1345 (0.0699, 0.2042) |
| | gte-Qwen2-1.5B-instruct (c2000, k20) | 184 | <0.001 | 1 | -0.0127 (-0.0572, 0.0328) | 0.3443 (0.2253, 0.4603) | 0.1945 (0.1295, 0.2648) |
| | Medical-T5-Large (c1000, k200) | 185 | <0.001 | 1 | -0.0099 (-0.0557, 0.0349) | 0.2170 (0.1188, 0.3240) | 0.1573 (0.0948, 0.2284) |

| | | | | | | | |
|---|---|---|---|---|---|---|---|
| | text-embedding-3-large (c2000, k200) | 183 | <0.001 | 1 | -0.0158 (-0.0616, 0.0287) | 0.2522 (0.1504, 0.3602) | 0.1661 (0.1099, 0.2268) |

* Sens. and Spec. Holm p, p-values from exact McNemar tests comparing sensitivity and specificity with PheNorm, adjusted using the Holm method across the 24 comparisons for each metric. Δ = RAG − PheNorm, with 95% CIs from 2,000 paired bootstrap resamples; CIs excluding zero indicate significant differences. n, number of test-set patients with RAG outputs; outputs were unavailable for some patients because the input exceeded the GPT-4o API's maximum input length or triggered the Azure OpenAI content filter. Both methods were compared on the same patients.
